# Clinical Features, Radiological Findings, and Outcome in Patients with Symptomatic Mild (<50%) Carotid Stenosis: A MUSIC Study

**DOI:** 10.1101/2024.05.02.24306793

**Authors:** Daina Kashiwazaki, Kohei Chida, Kazumichi Yoshida, Kiyofumi Yamada, Motohiro Morioka, Katsuhiko Maruichi, Emiko Hori, Naoki Akioka, Yasushi Takagi, Junta Moroi, Susumu Miyamoto, Toru Iwama, Masaki Chin, Kenji Kamiyama, Kojiro Wada, Nobuyuki Sakai, Tsuyoshi Izumo, Yusuke Nishikawa, Mitsuhito Mase, Kohkichi Hosoda, Katsumi Takizawa, Eiichi Kobayashi, Michiya Kubo, Atsushi Fujita, Taku Sugiyama, Miki Fujimura, Hideyuki Yoshioka, Hiroyuki Kinouchi, Takeharu Kunieda, Ataru Nishimura, Shinichi Yoshimura, Yoshiaki Shiokawa, Hiroshi Abe, Hiroharu Kataoka, Kuniaki Ogasawara, Masaaki Uno, Makoto Sasaki, Satoshi Kuroda, the MUSIC Study Group

**Affiliations:** Department of Neurosurgery, Graduate School of Medicine and Pharmaceutical Sciences, University of Toyama, Toyama, Japan; Department of Neurosurgery, School of Medicine, Iwate Medical University, Morioka, Japan; Department of Neurosurgery, Shiga University of Medical Science, Otsu, Japan; Department of Neurosurgery, National Cerebral and Cardiovascular Center, Suita, Japan; Department of Neurosurgery, Kurume University School of Medicine, Kurume, Japan; Department of Neurosurgery, Kashiwaba Neurosurgical Hospital, Hokkaido, Japan; Department of Neurosurgery, Tokushima University Graduate School of Biomedical Sciences, Tokushima, Japan; Department of Surgical Neurology, Akita Cerebrospinal and Cardiovascular Center, Akita, Japan; Department of Neurosurgery, Kyoto University Graduate School of Medicine, Kyoto, Japan; Department of Neurosurgery, Gifu University Graduate School of Medicine, Gifu, Japan; Department of Neurosurgery, Kurashiki Central Hospital, Kurashiki, Japan; Department of Neurosurgery, Nakamura Memorial Hospital, Sapporo, Japan; Department of Neurosurgery, National Defense Medical College Hospital, Tokorozawa, Japan; Department of Neurosurgery, Kobe City Medical Center General Hospital, Kobe Japan; Department of Neurosurgery, Nagasaki University Graduate School of Biomedical Sciences, Nagasaki, Japan; Department of Neurosurgery, Graduate School of Medical Science, Nagoya City University, Nagoya, Japan; Department of Neurosurgery, Kobe City Nishi-Kobe Medical Center, Kobe, Japan; Department of Neurosurgery, Asahikawa Red-Cross Hospital, Asahikawa, Japan; Department of Neurosurgery, National Hospital Organization Chiba Medical Center, Chiba, Japan; Department of Neurosurgery, Saiseikai Toyama hospital, Toyama, Japan; Department of Neurosurgery, Kobe University Graduate School of Medicine, Kobe, Japan; Department of Neurosurgery, Hokkaido University Graduate School of Medicine, Sapporo, Japan; Department of Neurosurgery, Interdisciplinary Graduate School of Medicine and Engineering, University of Yamanashi, Chuo, Japan; Department of Neurosurgery, Ehime University Graduate School of Medicine, Ehime, Japan; Department of Neurosurgery, Graduate School of Medicine, Kyushu University, Fukuoka, Japan; Department of Neurosurgery, Hyogo Medical University, Nishinomiya, Japan; Department of Neurosurgery, Fuji Brain Institute and Hospital, Fujinomiya, Japan; Department of Neurosurgery, Faculty of Medicine, Fukuoka University, Fukuoka, Japan; Department of Neurosurgery, Kawasaki Medical School, Okayama, Japan; Division of Ultrahigh Field MRI, Institute for Biomedical Sciences, Iwate Medical University, Iwate, Japan

**Keywords:** carotid endarterectomy, ischemia, mild carotid stenosis, stroke

## Abstract

**Background and Purpose:** Plaque composition, but not stenosis degree, may play a key role in the development of recurrent ischemic events in patients with symptomatic, mild (<50%) carotid stenosis. This multi-center prospective cohort study was aimed to determine their clinical and radiological features and to evaluate the benefits of carotid endarterectomy (CEA) for them.

**Methods:** This study included 124 patients with cerebrovascular or retinal ischemic events ipsilateral to mild carotid stenosis. Best medical therapy (BMT) was administered to all participants. CEA or carotid artery stenting (CAS) was implemented at each institution’s discretion. Baseline and 6-, 12-, and 24-month follow-up data were collected. Primary endpoint was ipsilateral ischemic stroke. Secondary endpoints included any stroke, ipsilateral TIA, ipsilateral ocular symptoms, any death, and plaque progression requiring CEA/CAS. Multivariate Cox proportional hazard model was used to evaluate the predictors for each endpoint.

**Results:** Of 124 patients, 70 patients (56.5%) had the history of ipsilateral ischemic stroke and 51 (43.5%) had been treated with antiplatelet agents. Mean stenosis degree was 22.4±13.7%. Plaque composition was categorized into fibrous plaque in 22 patients, lipid-rich/necrotic core (LR/NC) in 25, and intraplaque hemorrhage (IPH) in 69. BMT was indicated for 59 patients, while CEA was performed in 63. The incidence of primary endpoint was significantly higher in BMT group than in CEA group (15.1% vs. 1.7%; HR, 0.18; 95%CI, 0.05–0.84; P=0.03). The predictors for ipsilateral ischemic stroke were CEA (HR, 0.18; 95%CI, 0.05–0.84; P=0.03) and IPH (HR, 1.92; 95%CI, 1.26–4.28; P=0.04). The results on secondary endpoints were very similar.

**Conclusion:** IPH may highly predict subsequent cerebrovascular events, whereas CEA may reduce these risk during a 2-year follow-up in patients with symptomatic, mild carotid stenosis.

**Registration:** This study has been registered in the University Hospital Medical Information Network Clinical Trials Registry (UMIN-CTR) (UMIN000023635).

**Contributors:** SK, KO, UM, and MS designed the study. DK wrote an original draft. All authors recruited patients. DK, KC, KY, KY, MM, SY, SM, KO, MU, MS, and SK analyzed the imaging data as the members of Central Judgement Committee and contributed to data analysis. SK takes final responsibility for the submitted publication.

---

Ischemic stroke is a leading cause of death and long-term disability worldwide.^1-3^ Atherosclerotic carotid artery stenosis is one of most important risk factors for ischemic stroke, contributing to up to 20–30% of all cases of ischemic stroke or TIA.^4^ The degree of carotid stenosis has long been used to estimate the risk of recurrence of ischemic events and to stratify treatment indications. For these three decades, carotid endarterectomy (CEA) is believed to yield no significant benefits for patients with mild (<50%) stenosis of carotid artery.^3^ With advances in imaging technology, however, there is an increasing evidence that a certain subgroup of medically treated patients with mild carotid stenosis may have at higher risk to repeat ischemic stroke when their plaque is vulnerable.^5-7^ Thus, it may be no further sufficient to assess the stenosis degree alone to predict the future risk of ischemic events in patients with symptomatic mild carotid stenosis^5^.

Therefore, this multi-center prospective cohort study, “Mild but Unstable Stenosis of Carotid Artery (MUSIC) Study” was aimed to characterize clinical and radiological features and outcome in patients with symptomatic mild carotid stenosis. Especially, we precisely analyzed the beneficial effects of CEA on their incidence of recurrent ischemic events during a 2-year follow-up.

## Methods

### Data availability statement

The data analyzed in the current study are available from the corresponding author on reasonable request.

### Ethical aspects

This study was conducted according to Good Clinical Practice guidelines and the Declaration of Helsinki, and STROBE statement. This study was approved by Institutional Review Board at each institute. All participants provided written informed consent when enrolled in this study.

### Study design

For this multi-center, prospective cohort study, the participants were recruited from 28 hospitals in Japan. Between January 2017 and December 2020, the patients who meet all of the following criteria were included in this study: (1) their age was 20 years or older, (2) they experienced TIA, ischemic stroke, amaurosis fugax, or retinal artery occlusion within 6 months of enrollment, (3) they had mild (<50%) cervical internal carotid artery stenosis on the ipsilateral side as the above, (4) their daily living was independent (modified Rankin scale [mRS], 0-2), (5) they could visit outpatient clinic for the 2-year follow-up, and (6) their written consent to participate in this study was obtained. The degree of internal carotid stenosis was determined according to the North American Symptomatic Carotid Endarterectomy Trial (NASCET) criteria^3^. The exclusion criteria were as follows: the patients with asymptomatic carotid stenosis, the patients with symptomatic, but moderate to severe (≥50%) carotid artery stenosis; contraindications to magnetic resonance imaging (MRI); carotid artery dissection; or competing etiologies. Simultaneously, we collected the number of patients who were admitted to the participating hospitals due to symptomatic carotid stenosis during the 4-year enrollment.

Following information were collected at the enrollment: age, gender, clinical diagnosis, onset date, neurological symptoms on admission, comorbidities, family history, past history, medications, mRS score, blood pressure, and laboratory data. Following radiological data were also collected: brain MRI (FLAIR, and T1-, T2-, and diffusion-weighted images), brain MRA, neck MRI (T1-weighted images), neck MRA, and 3D-CTA or cerebral angiography.

### Treatments and follow-up

The BMT, including the antiplatelets and the agents for hypertension, hyperlipidemia, and diabetes mellitus, was administered to all participants as appropriate. All current smokers were instructed to cease smoking immediately. In addition to these medical treatments, CEA or CAS was performed at the discretion of each hospital.

All participants were followed up at each outpatient clinic for two years. Neurological status, laboratory data, and blood pressure were assessed at baseline and 6, 12, and 24 months of follow-up. When pathological values were observed, appropriate treatment was initiated to maintain target low-density lipoprotein (LDL)-cholesterol values of <140 mg/dL and blood pressure of ≤140/90 mmHg. In this study, the temporal profile of mRS score, blood pressure, and LDL-cholesterol were analyzed at 6, 12, and 24 months of follow-up.

### Assessment of imaging studies

After the 2-year follow-up, a Central Judgement Committee performed blind evaluations of images. All committee members were the physicians who were specialized in the diagnosis and treatment of carotid artery stenosis (DK, KC, KY, KY, MM, SY, KO, MU, MS, SK).

Brain MRI was used to localize cerebral infarction that caused the current ischemic events. Neck MRI was used to determine the composition of plaques. The signal intensity ratio (SIR) was calculated as the ratio of the signal intensity of carotid plaque to that of the adjacent sternocleidomastoid muscle on T1-weighted images. According to previous report, when the SIR value was less than 1.2 (iso-intensity), the plaque was considered fibrous; when the SIR value was between 1.2 and 1.5 (relatively high intensity), the plaque was considered lipid-rich or necrotic core (LR/NC). If the SIR value was greater than 1.5 (high intensity), the plaque was considered to be mainly composed of intraplaque hemorrhage (IPH).^8^ Cerebral angiography or 3D-CTA was used to determine the degree of stenosis and to detect carotid webs and ulcer formation in the plaque.

All images were independently evaluated by two members of Central Judgement Committee to determine the inter-observer reproducibility. In addition, each reader reviewed the radiological findings twice with an interval of 4 weeks, blinded to the first reading, to determine the intra-rater reproducibility. Finally, the agreements within and between observers were quantified, using the linear weighted κ statistic or intraclass correlation coefficient (ICC).^9^

### Endpoints

In this study, the primary endpoint was set as the occurrence of ipsilateral cerebral ischemic stroke during a 2-year follow-up. The secondary endpoints included the occurrence of any stroke, ipsilateral TIA, ipsilateral ocular symptoms, any death, and plaque progression requiring CEA/CAS.

### Statistical analysis

Continuous variables were expressed as the mean ± standard deviation (SD) or the median and interquartile range (IQR). For statistical comparisons between groups. clinical characteristics were compared using *t*-test, Mann-Whitney U test, χ^2^ test, or one-factor ANOVA as appropriate. Cumulative incidence of endpoints was analyzed using the Kaplan-Meier method. For the occurrence of the endpoints, Cox proportional hazards regression model was used to estimate the hazard ratios (HRs) and 95% confidence intervals (CIs). We also used the Cochran-Mantel-Haenszel test to compare the distribution of the 2-years mRS score between two groups. Differences were considered statistically significant at P<0.05. Missing data completion was not performed. Complete-case analysis was performed when missing data were included.

## Results

### Patients

This study enrolled 124 patients during 4 years between January 2017 and December 2020. During this period, 1,208 patients with symptomatic carotid stenosis were treated in the participating hospitals. Therefore, 124/1,208 (about 10%) of symptomatic carotid stenosis patients had mild (<50%) carotid stenosis.

Of these 124 patients, there were 111 males (89.5%) and 13 females (10.5%). Their mean age was 74.4±8.6 years. Clinical diagnosis at the enrollment were TIA (n=10), ischemic stroke (n=98), and ocular symptoms (n=16). Past history included hypertension in 90 patients (72.6%), diabetes mellitus in 43 (34.7%), and dyslipidemia in 69 (55.5%). Mean systolic and diastolic blood pressure at the enrollment was 136.9±21.4 and 75.6±12.2 mmHg, respectively. Mean value of LDL cholesterol was 98.6±32.0 mg/dL.

A significant number of patients had cardiovascular diseases prior to the cerebrovascular or ocular event that led to their enrollment in this study, including coronary artery disease in 31 patients (25.0%), aortic disease in 15 (12.1%), ASO in 5 (4.0%), and chronic renal failure in 14 (11.3%). In addition, 70 patients (56.5%) and 7 patients (5.6%) had had the history of ischemic and hemorrhagic stroke, respectively. About half of the patients (59/124, 47.6%) had a history of ischemic stroke on the ipsilateral side of the mild carotid stenosis that led to their enrollment into this study. Of these, 12 patients had repeated ischemic stroke on the ipsilateral side before being enrolled in the study despite the BMT in each hospital: the number of episodes of ipsilateral ischemic stroke was two (n=8), three (n=3), and four (n=1). In spite of single (n=43) or dual antiplatelet therapy (n=11), cerebrovascular or ocular event occurred and led to their enrollment into this study. Other 8 patients (6.5%) had been treated with anticoagulants. Therefore, a total of 62 patients (50%) experienced the cerebrovascular or ocular event that led to their enrollment into this study despite their prior antithrombotic therapy.

### Radiological findings

Mean stenosis degree was 22.4±13.7% according to the NASCET criteria. The stenosis degree was judged as 0% in 20 patients (16.1%). Ulceration was identified in 60 patients (48.4%). Carotid web was found in 4 patients (3.2%). **Fig. 1A** shows plaque composition at the enrollment in 116/124 patients. Plaque images in other 8 patients were excluded because of poor image quality. Of 116 patients, 22 (19.0%) had fibrous plaque and 25 (21.5%) had LR/NC plaque. The remaining 69 patients (59.5%) had IPH. Therefore, 94 patients (81%) had radiologically unstable plaque (LR/NC + IPH).

**Fig. 1.**
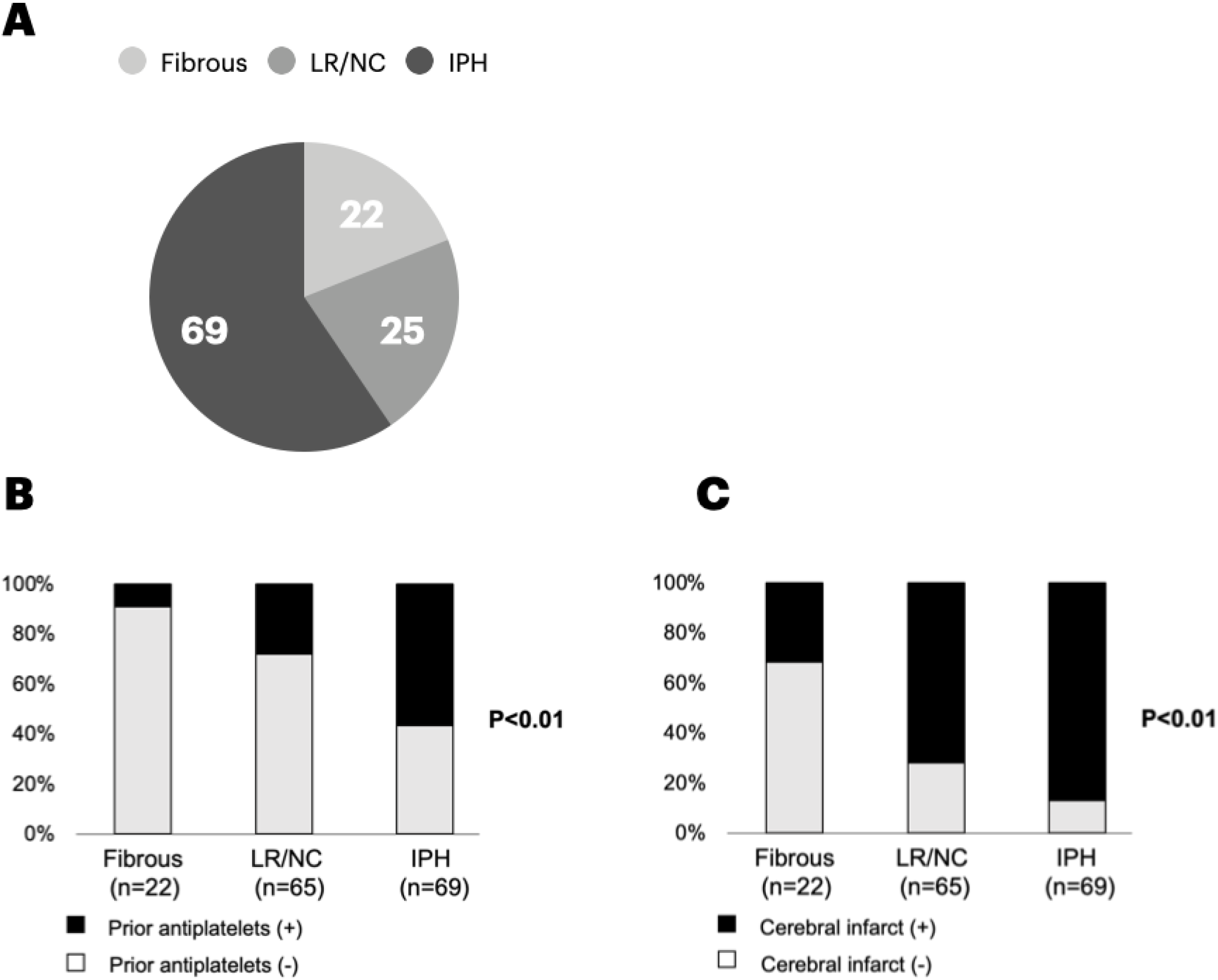
Clinical and radiological characteristics in patients with symptomatic mild carotid stenosis. (A) Plaque composition on MRI. (B) The prevalence of prior antiplatelet therapy in patients with fibrous plaque, LR/NC, and IPH. (C) The prevalence of cerebral infarct in patients with fibrous plaque, LR/NC, and IPH.

As shown in **Fig. 1B**, antiplatelet therapy had been performed in 39/69 (56.5%) patients with IPH prior to the cerebrovascular or ocular event that led to their enrollment into this study, being significantly higher than in those with fibrous plaque (2/20, 10%) and in those with lipid-rich plaque (7/25, 28%; P<0.01). There was no correlation between the stenosis degree and plaque composition (**Fig. S1**).

On diffusion-weighted MRI, cerebral infarct was found in 87/124 patients (70.2%). Cerebral infarct was located in the cortex in 29 patients (33.3%), the white matter in 14 (16.1%), or both in 44 (50.6%). Cerebral infarct was found in 7/22 patients with fibrous plaque (31.8%), 18/25 patients with LR/NC (72.0%), and 60/69 patients with IPH (87.0%). Therefore, the prevalence of cerebral infarct was significantly higher in patients with LR/NC or IPH than in those with fibrous plaque (P<0.001; **Fig. 1C**).

**Table S1** shows the inter-observer and intra-observer reproducibility about the stenosis degree, ulceration, carotid web, cerebral infarct presence, cerebral infarct location, SIR, plaque composition, and IPH presence.

### Treatments and follow-up

As shown in **Fig. 2**, 59 patients (47.6%) were medically treated (BMT group). Other 63 patients (50.8%) underwent CEA and two patients (1.6%) underwent CAS as well as BMT. Two-year follow-up was completed in 51/59 patients in BMT group, 59/63 patients in CEA group, and 2/2 patients in CAS group (n=112). The remaining 12 patients (9.7%) were lost during a 2-year follow-up. Therefore, we compared the incidence of primary and secondary endpoints between BMT and CEA groups, because the sample size of CAS group was too small for statistical analysis (n=2).

**Fig. 2.**
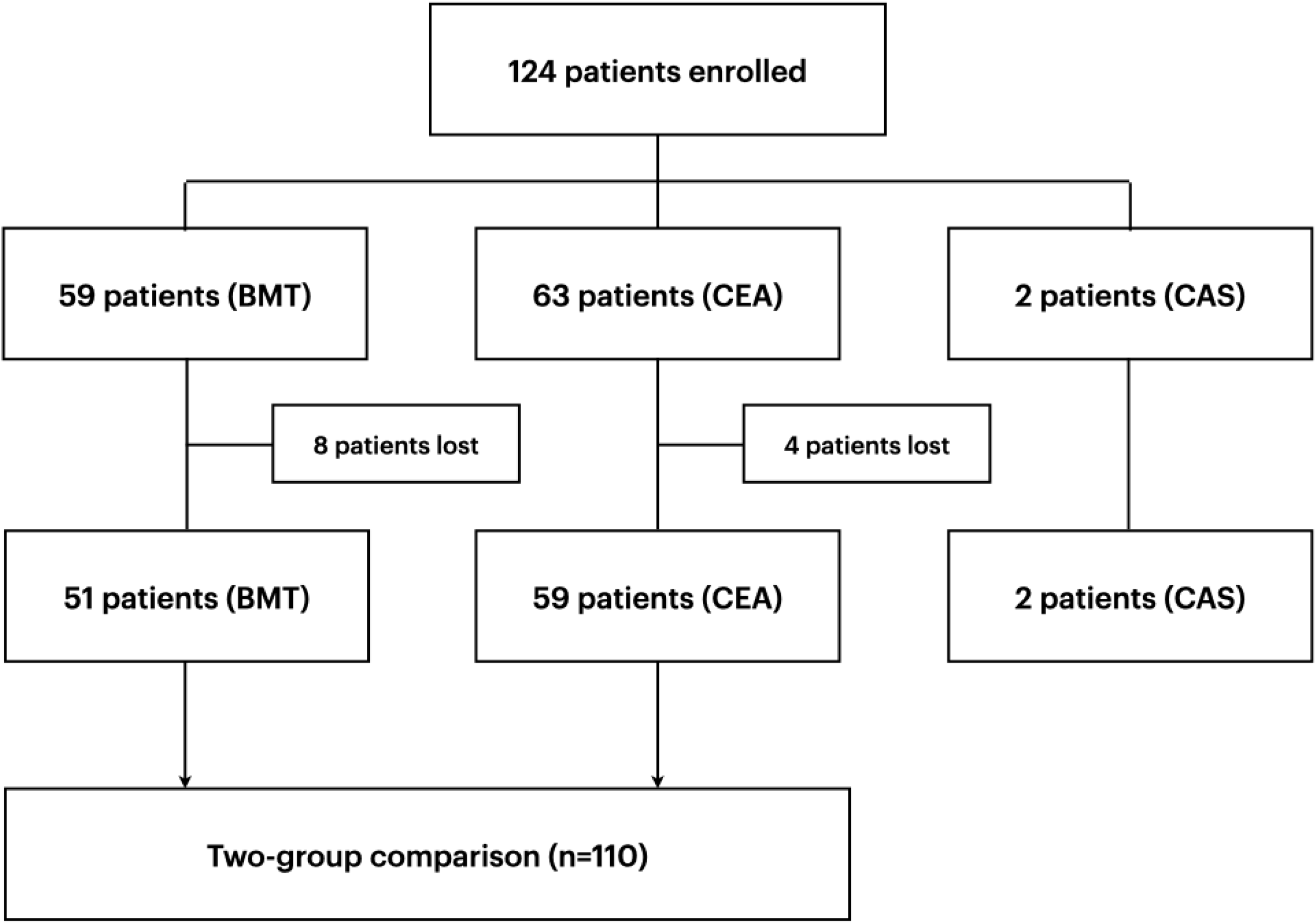
Flow chart showing the patient selection process. BMT, best medical treatment; CEA, carotid endarterectomy; CAS, carotid artery stenting

**Table 1** demonstrates the clinical and radiological characteristics in BMT and CEA groups. There were no significant differences in their clinical and radiological data except for the history of ipsilateral ischemic stroke, of which incidence was significantly higher in CEA group (36/59, 61%) than in BMT group (18/51, 35.3%; P=0.003). Blood pressure and LDL-cholesterol value were appropriately controlled in a majority of patients. There was no significant difference in systolic and diastolic blood pressure during a 2-year follow-up between two groups **(Fig. S2)**. No significant difference in LDL-cholesterol value was observed during a 2-year follow-up between two groups **(Fig. S3)**.

**Table 1.** Clinical characteristics at the enrollment in BMT and CEA groups. BMT, best medical treatment; CEA, carotid endarterectomy; ASO, arteriosclerosis obliterans;

| Variables | BMT (n=51) | CEA (n=59) | P |
| --- | --- | --- | --- |
| Age (years) | 74.9±8.2 | 74.0±8.5 | 0.65 |
| Gender |  |  |  |
| male | 44 | 53 | 0.77 |
| female | 7 | 6 |  |
| Medical comorbidity |  |  |  |
| Hypertension | 37 | 44 | 0.49 |
| Diabetes | 14 | 20 | 0.41 |
| Hypercholesterolemia | 26 | 35 | 0.24 |
| Coronary artery disease | 9 | 18 | 0.12 |
| Aortic disease | 7 | 6 | 0.77 |
| ASO | 2 | 4 | 0.37 |
| Chronic renal failure | 5 | 7 | 0.76 |
| History of stroke |  |  |  |
| Ipsilateral ischemic stroke | 18 | 36 | 0.003 |
| Other ischemic strokes | 4 | 6 | 0.75 |
| Hemorrhagic stroke | 2 | 2 | 0.88 |
| Systolic blood pressure(mmHg) | 138.6±19.2 | 138.6±19.0 | 0.88 |
| Diastolic blood pressure(mmHg) | 76.4±10.5 | 75.1±10.2 | 0.56 |
| LDL cholesterol (mg/dL) | 101.3±33.6 | 95.8±32.5 | 0.28 |
| Medical therapy prior to enrollment |  |  |  |
| Antihypertensives | 38 | 43 | 0.7 |
| Statins for HL | 21 | 24 | 0.85 |
| Others for HL | 4 | 4 | 0.97 |
| Anti-DM agents | 19 | 15 | 0.17 |
| Antiplatelets | 20 | 30 | 0.1 |
| anticoagulants | 4 | 3 | 0.97 |
| mRS score |  |  |  |
| 0 | 22 | 14 | 0.22 |
| 1 | 17 | 30 |  |
| 2 | 10 | 12 |  |
| Radiological findings |  |  |  |
| Degree of stenosis (%) | 22.9 | 20.5 | 0.36 |
| Plaque ulceration presence | 25 | 35 | 0.17 |
| Plaque composition |  |  |  |
| fibrous | 11 | 7 | 0.32 |
| LR/NC | 12 | 11 |  |
| IPH | 27 | 37 |  |

**Table 2** demonstrates the occurrence of primary endpoint during a 2-year follow-up in two groups. Ipsilateral ischemic stroke occurred in 8/51 BMT patients (15.7%) but occurred in 1/59 CEA patients (1.7%) during CEA procedure. None of CEA patients experienced it during a 2-year follow-up. As shown in **Fig. 3A**, multivariate Cox proportional hazards model demonstrated that the incidence of ipsilateral ischemic stroke was significantly lower in CEA group than in BMT group (HR, 0.18; 95%CI, 0.05–0.84; P=0.03). As shown in **Table 3**, the predictors for ipsilateral ischemic stroke were CEA (HR, 0.18; 95%CI, 0.05–0.84; P=0.03) and IPH (HR, 1.92; 95%CI, 1.26–4.28; P=0.04).

**Table 2.** The incidence of patients experiencing primary/secondary endpoints during a 2-year follow-up in BMT and CEA groups. BMT, best medical treatment; CEA, carotid endarterectomy; TIA, transient ischemic attack; CAS, carotid artery stenting The number in parentheses represents the number of patients with intraplaque hemorrhage.

|  | BMT group (n=51) | CEA group (n=59) |
| --- | --- | --- |
| **Primary endpoint** |  |  |
| Ipsilateral ischemic stroke | 8 (7) | 0 (0) |
| Perioperative ischemic stroke |  | 1 (1) |
| **Secondary endpoint** |  |  |
| any ischemic stroke | 9 (8) | 1 (1) |
| ipsilateral TIA | 1 (1) | 1 (0) |
| ipsilateral ocular symptoms | 1 (0) | 0 (0) |
| any hemorrhagic stroke | 0 (0) | 0 (0) |
| any death | 2 (1) | 2 (1) |
| stenosis progression requiring CEA/CAS | 3 (3) | 0 (0) |

**Table 3.** Predictors for primary and secondary endpoints. ASO, arteriosclerosis obliterans; IPH, intraplaque hemorrhage; HR, hazard ratio; 95%CI, 95% confidential interval

| Variables | Primary endpoint |  |  |  | Secondary endpoint |  |  |  |
| --- | --- | --- | --- | --- | --- | --- | --- | --- |
|  | Univariate |  | Multivariate |  | Univariable |  | Multivariable |  |
|  | HR (95%CI) | P | HR (95%CI) | P | HR (95%CI) | P | HR (95%CI) | P |
| Age ($\geq 75$ years) | 1.09 (0.82-1.28) | 0.78 | | | 1.14 (0.87-1.39) | 0.28 | | |
| Gender (male) | 1.03 (0.86-1.56) | 0.64 |  |  | 1.06 (0.88-1.44) | 0.42 |  |  |
| Carotid endarterectomy (yes) | 0.11 (0.03-0.81) | 0.03 | **0.18 (0.05-0.84)** | **0.03** | 0.26 (0.11-0.65) | 0.005 | **0.32 (0.13-0.79)** | **0.01** |
| Hypertension (yes) | 1.15 (0.91-1.41) | 0.23 |  |  | 1.13 (0.88-1.42) | 0.28 |  |  |
| Diabetics (yes) | 0.98 (0.70-1.38) | 0.89 |  |  | 1.05 (0.89-1.50) | 0.57 |  |  |
| Dyslipidemia (yes) | 1.05 (0.88-1.40) | 0.55 |  |  | 1.04 (0.87-1.46) | 0.51 |  |  |
| Coronary disease (yes) | 1.11 (0.81-1.50) | 0.34 |  |  | 1.14 (0.81-1.48) | 0.24 |  |  |
| Aortic disease (yes) | 1.08(0.83-1.26) | 0.65 |  |  | 1.04 (0.89-1.19) | 0.75 |  |  |
| ASO | 1.12 (0.86-1.35) | 0.21 |  |  | 1.12 (0.86-1.35) | 0.21 |  |  |
| Renal failure (yes) | 0.96 (0.91-1.04) | 0.48 |  |  | 1.15 (0.85-1.26) | 0.19 |  |  |
| Past ischemic stroke (yes) | 1.10 (0.90-1.37) | 0.26 |  |  | 1.13 (0.88-1.23) | 0.28 |  |  |
| Past ipsilateral ischemic stroke (yes) | 1.14 (0.94-1.44) | 0.18 |  |  | 1.12 (0.81-1.21) | 0.3 |  |  |
| Stenosis degree ($\geq 22\%$) | 0.96 (0.65-1.34) | 0.71 | | | 0.94 (0.61-1.45) | 0.43 | | |
| IPH presence (yes) | 1.60 (1.08-3.43) | 0.04 | **1.92 (1.26-4.28)** | **0.04** | 1.50 (1.07-4.21) | 0.03 | **1.52 (1.06-4.39)** | **0.03** |
| Ulceration presence (yes) | 1.12 (0.89-1.39) | 0.2 |  |  | 1.08 (0.83-1.29) | 0.38 |  |  |
| Cerebral infarction presence (yes) | 1.06 (0.81-1.49) | 0.28 |  |  | 1.05 (0.89-1.42) | 0.25 |  |  |

**Fig. 3.**
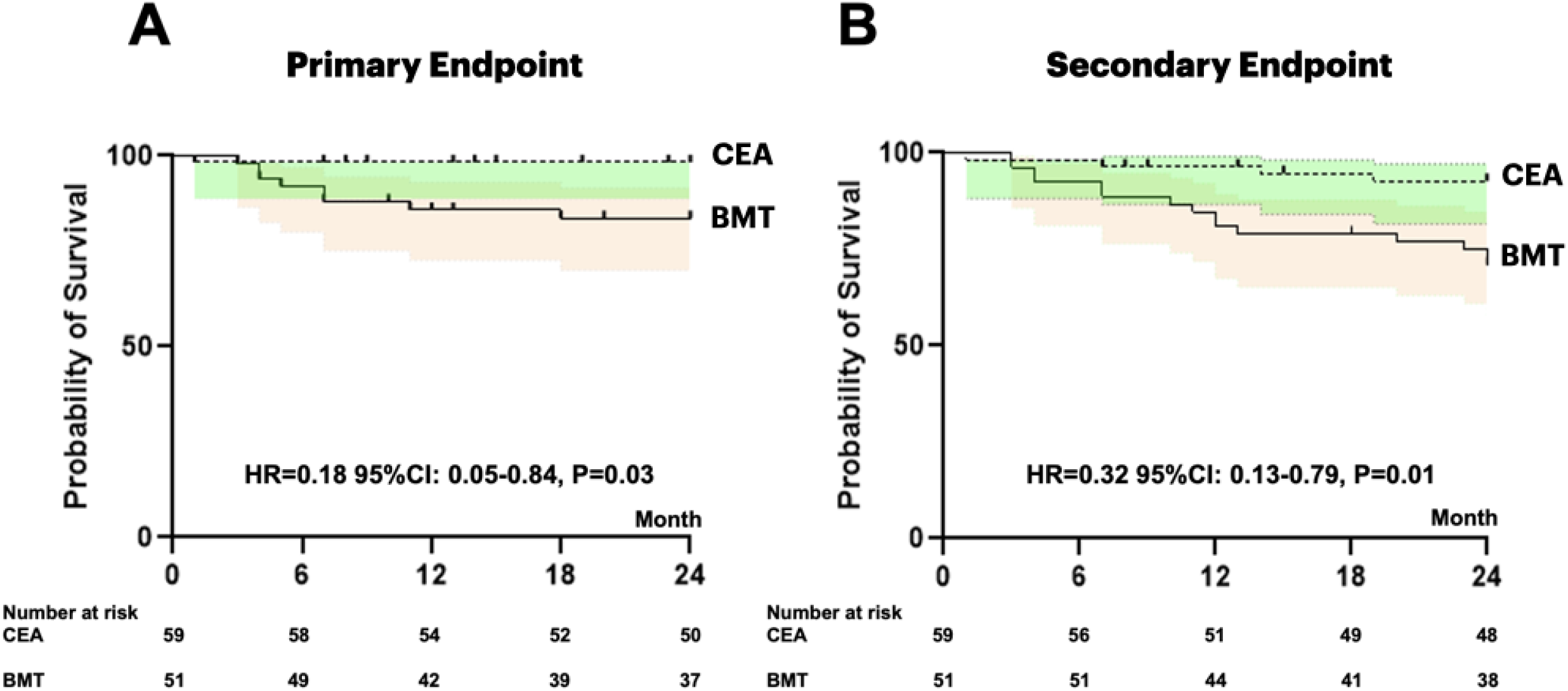
Kaplan-Meier survival curves. (A) The incidence of primary endpoint was significantly lower in CEA group than in BMT group (HR=0.11 [95% CI:0.03-0.81], P=0.03). (B) The incidence of secondary endpoints was significantly lower in CEA group than in BMT group (HR=0.26 [95% CI:0.11-0.65], P=0.005). BMT, best medical treatment; CEA, carotid endarterectomy; HR, hazard ratio; 95%CI, 95% confidence interval.

**Table 2** demonstrates the occurrence of secondary endpoints during a 2-year follow-up in two groups. Secondary endpoints occurred in 16/51 BMT patients (31.4%), including any ischemic stroke (n=9), ipsilateral TIA (n=1), ipsilateral ocular event (n=1), any death (n=2), and stenosis progression requiring CEA/CAS (n=3). However, secondary endpoints occurred in 4/55 CEA patients (7.3%), including any ischemic stroke (n=1), ipsilateral TIA (n=1), and any death (n=2). As shown in **Fig. 3B**, multivariate Cox proportional hazards model demonstrated that the incidence of secondary endpoint was significantly lower in CEA group than in BMT group (HR, 0.32; 95%CI, 0.13–0.79; P=0.01). As shown in **Table 3**, the predictors for secondary endpoints were CEA (HR, 0.32; 95%CI, 0.13–0.79; P=0.01) and IPH (HR, 1.52; 95%CI, 1.06–4.39; P=0.03).

**Fig. S4** shows the mRS scores at the enrollment and 2-year follow-up. There was no significant difference at the enrollment between two groups. However, the prevalence of favorable functional outcome (mRS, 0–2) at 2 years were significantly higher in CEA group (50/52, 96.3%) than in BMT group (43/50, 86.0%; P<0.01). A favorable shift in mRS score was also observed in CEA group, but not in BMT group (odds ratio [OR], 2.4; 95%CI, 1.3-4.0; P=0.02).

## Discussion

This is the first multi-center prospective cohort study to evaluate clinical and radiological features in symptomatic patients with mild (<50%) carotid stenosis. In addition, this study is very unique to compare their 2-year outcomes between BMT and CEA groups. This study mainly yields three novel findings, including clinical features, radiological findings, and treatment outcome in symptomatic, mild carotid stenosis.

### Clinical features

The subjects who fulfilled the eligibility was about 10% of patients who were treated at each participating institute for symptomatic carotid stenosis. Therefore, the patients with symptomatic mild carotid stenosis are not rare in the real world. The subjects were frequently associated with vascular comorbidities, including coronary artery disease (25%), aortic disease (12%), and chronic renal failure (11%). About 50% of them had the history of ischemic stroke ipsilateral to mild carotid stenosis. Therefore, the subjects included in this study are highly complicated by a variety of comorbidities even if the stenosis degree was mild. More interestingly, about a half of them had been treated with antiplatelets or anticoagulants prior to the enrollment in this study, which strongly suggests that a certain subgroup of patients with symptomatic mild carotid stenosis are refractory to medical therapy. Therefore, they should not be prematurely assumed to be at low risk for subsequent cerebrovascular events, only because the stenosis degree is mild.

### Radiological findings

In this cohort, the stenosis degree was judged as “0%” in about 15% of subjects (20/124). One of the reasons for the very low stenosis despite the presence of plaque with significant volume may be the positive remodeling that commonly occurs in the atherosclerotic arteries. Positive remodeling is a phenomenon in which the diseased arteries dilate their outer diameter with the formation of plaque. Macrophage-producing proteinase may play an important role in this phenomenon. ^10, 11^ Recent studies have shown that positive remodeling is more common in symptomatic t patients. ^12^ It is closely related to unstable plaque and infiltration of macrophages.^13, 14^ It also correlates with diffusion MRI-positive lesions and ischemic events after CEA and CAS. ^15, 16^ Based on these findings, the information on plaque volume and composition on MRI is now essential when evaluating patients with mild carotid stenosis as well as the information on luminal morphology on conventional modalities such as cerebral angiography.

This study demonstrates a very high prevalence of unstable plaques, including LR/NC (22%) and IPH (60%). There are few studies that precisely denote the plaque composition in mild carotid stenosis, but recent widespread use of MRI has gradually clarified it. Thus, these unstable plaque can be confirmed in all 18 patients who underwent CEA for recurrent symptomatic mild carotid stenosis.^17^ IPH and positive remodeling are found in about 60% of patients with symptomatic, mild carotid stenosis.^18^ Unstable plaque can be found in 10/11 patients who underwent CEA for symptomatic mild carotid stenosis.^19^ Therefore, a majority of patients with symptomatic, mild carotid stenosis should be recognized as having unstable plaques.

Many patients with IPH repeated ischemic events ipsilateral to the mild carotid stenosis regardless of antiplatelet therapy. In fact, 4 of 22 patients with symptomatic mild carotid stenosis experienced TIA or stroke recurrence despite antiplatelet therapy.^20^ Therefore, antiplatelet therapy can of course reduce the risk of subsequent cerebrovascular events in most of these patients, but the efficacy may largely be limited in a certain subgroup of patients with IPH. ^21-23^ A distinctive clinical and radiological features in these high-risk patients strongly indicate that plaque composition, but not stenosis degree, is playing a key role for subsequent ischemic events in patients with symptomatic, mild carotid stenosis. ^5^

### Treatment outcome

Current guidelines have not recommended CEA for patients with symptomatic, mild carotid stenosis.^24, 25^ In this study, however, CEA significantly reduced the incidence of both primary and secondary endpoints during a 2-year follow-up. In fact, Yoshida et al. (2012) medically followed up 25 patients with symptomatic mild carotid stenosis due to unstable plaque and found that 46% of them developed recurrent ischemic events (annual risk, 17.8%).^26^ Kurosaki et al. (2022) also reported that about 40% of patients with mild carotid stenosis experienced recurrent ischemic events (annual risk, 8.4%).^18^ Recent meta-analysis has analyzed the risk for ipsilateral ischemic stroke and any operative stroke or death in patients with mild carotid stenosis. As the results, the annual risk in medically treated patients was 3.0% in patients with 30-49% stenosis and 1.8% in those with <30% stenosis. On the other hand, the meta-analysis found that the annual risk in surgically treated patients was 2.6% in patients with 30-49% stenosis and 2.2% in those with <30% stenosis.^27^ The risk in surgically treated group was much higher than our results. The reasons of this large discrepancy are obscure, but the controls of blood pressure and LDL-cholesterol during the 2-year follow-up period were favorable in this study, indicating that the quality of medical care in addition to CEA was adequate in our cohort.

This is the first study that denotes the predictors for the occurrence of primary and secondary endpoints in patients with symptomatic mild carotid stenosis. CEA significantly reduced their incidence, but IPH significantly increased it. Of course, this study is not a randomized clinical trial, thus we should be careful to consider the findings. But, clinical and radiological characteristics were very similar between BMT and CEA groups. In fact, there are several single-center studies that report the efficacy of CEA in patients with symptomatic mild carotid stenosis.^14, 17, 18, 28, 29^ On the other hands, unstable plaque has recently been recognized as a potential biomarker to represent systemic inflammation and thus predict subsequent cardiovascular events.^30-32^ In very near future, therefore, the evaluation of plaque composition would be an essential examination to predict the risk of further events and to determine treatment strategies in each patient with symptomatic mild carotid stenosis.

There are several limitations in this study. This is a non-randomized study, thus may carries a potential bias in surgical indication. To address this limitation, further randomized studies are warranted to explore the benefits of CEA for patients with symptomatic mild carotid stenosis. This study only analyzed the results during 2 years of follow-up. Longer follow-up periods would allow us to determine the outcome and effects of CEA in these populations.

In conclusion, a majority of patients with symptomatic mild carotid stenosis was associated with a variety of comorbidities, including ipsilateral ischemic stroke. A significant group of them are refractory to antiplatelet therapy. IPH would highly predict subsequent cerebrovascular events, whereas CEA significantly reduce their risk during a 2-year follow-up. Randomized clinical trials with plaque imaging as a surrogate marker may be warranted to further validate the efficacy of CEA in these patients.

## Non-standard Abbreviations and Acronyms

BMT: best medical therapy;
CAS: carotid artery stenting;
CEA: carotid endarterectomy;
CI: confidence interval;
DWI: diffusion-weighted imaging;
ICC: intraclass correlation coefficient;
FLAIR: fluid-attenuated inversion recovery;
HR: hazard ratio;
IPH: intraplaque hemorrhage;
IQR: interquartile range;
LDL: low-density lipoprotein;
LR/NC: lipid-rich or necrotic core;
MRI: magnetic resonance imaging;
mRS: modified Rankin Scale;
NASCET: North American Symptomatic Carotid Endarterectomy Trial;
SD: standard deviation;
SIR: signal intensity ratio;
OR: odds ratio;
T1WI: T1-weighted imaging;
T2WI: T2-weighted imaging;
TIA: transient ischemic attack;
TOF: time-of-flight

## Sources of Funding

none

## Disclosures

none

## Supplemental Materials

Tables S1 Figures S1–S4

## Appendix

Yusuke Egashira, MD,

Department of Neurosurgery, Gifu University Graduate School of Medicine

Hirotoshi Imamura, Masanori Goto, Ryu Fukumitsu, Yoshihiro Omura, Nobuyuki Ohara,

Department of Neurosurgery, Kobe City Nishi-Kobe Medical Center, Kobe, Japan

Hidehito Kimura,

Department of Neurosurgery, Kobe University Graduate School of Medicine, Kobe, Japan

Takai Hiroki,

Department of Neurosurgery, Kawasaki Medical School, Okayama, Japan

Yoichi Morofuji,

Department of Neurosurgery, Nagasaki University Graduate School of Biomedical Sciences, Nagasaki, Japan

Yoshitaka Kurosaki,

Department of Neurosurgery, Kurashiki Central Hospital, Kurashiki, Japan

Masaaki Korai

Department of Neurosurgery, Tokushima University Graduate School of Biomedical Sciences, Tokushima, Japan

Hiromasa Kobayashi,

Department of Neurosurgery, Faculty of Medicine, Fukuoka University, Fukuoka, Japan

Yasuyuki Yoshida,

Department of Surgical Neurology, Akita Cerebrospinal and Cardiovascular Center, Akita, Japan

Masakazu Okawa,

Department of Neurosurgery, Kyoto University Graduate School of Medicine, Kyoto, Japan

Shirabe Matsumoto,

Department of Neurosurgery, Ehime University Graduate School of Medicine, Ehime, Japan

## Notes

### Competing Interest Statement

The authors have declared no competing interest.

### Clinical Trial

University Hospital Medical Information Network Clinical Trials Registry (UMIN-CTR) (registration number: UMIN000023635)

### Funding Statement

No funding

### Author Declarations

Ethical Review Board, Toyama University Hospital (No. R2016042)

